# Monocyte-to-lymphocyte and Platelet-to-lymphocyte ratios in Neuromyelitis Optica Spectrum Disorders: systematic review and meta-analysis

**DOI:** 10.64898/2026.11.12.26367908

**Authors:** Abdurrahman Elsayed Shouman, Asaad Ibrahim Abdalla, Reem Saad, Mahmoud Sherif, Mohammad Bdour, Hossam Rostom

## Abstract

**Background:** Neuromyelitis Optica Spectrum Disorders (NMOSD) are a group of neuroinflammatory conditions that affect the nervous system. MLR and PLR are simple inflammatory biomarkers and are studied in many autoimmune and demyelinating diseases. In this study, we investigate the available evidence to determine whether MLR and PLR are higher in NMOSD patients than in healthy subjects.

**Methods:** Systematic review and meta-analysis of studies measuring levels of PLR and/or MLR in both NMOSD patients and healthy controls were conducted. Mean difference (MD) and 95% confidence interval (CI) were used to measure the difference in levels of MLR and PLR between patients and healthy subjects.

**Results:** For MLR, the main meta-analysis yielded a pooled MD of 0.125 [-0.114; 0.363]. I^2^ was 83.8%. Subgroup meta-analysis of age- and sex-matched studies, demonstrated a pooled MD of 0.066 [0.012; 0.12]. For PLR, meta-analysis revealed a pooled MD of 23.99 [6.03; 41.96]. I^2^ was 82.3%. Subgroup analysis for age- and sex-matched studies for PLR, showed a pooled MD of 14 [-19.68; 47.68].

**Conclusion:** The main analysis suggests that MLR is not higher in NMOSD patients than in healthy controls. However, subgroup analysis of age- and sex-matched studies and leave-one-out analysis excluding the outlier study showed a statistically significant MLR increase in patients. PLR was found to exhibit a modest increase in NMOSD patients compared with healthy controls. Nonetheless, subgroup analysis of age- and sex-matched studies showed a statistically non-significant increase. Results should be interpreted cautiously. The available evidence is inconsistent and further research is needed.

## 1. Introduction

‘Neuromyelitis Optica Spectrum Disorders’ (NMOSD) is an umbrella term that refers to a group of neuroinflammatory conditions that mainly affect the optic nerve, the spinal cord, and, less commonly, the brainstem^1^. Traditionally, Neuromyelitis Optica (NMO) referred to a neuroinflammatory condition where patients typically had optic neuritis and longitudinally extensive transverse myelitis (LETM)^2^. In 2005, an immunoglobulin G (IgG) antibody to the most abundant water channel in the brain (Aquaporin 4) was identified, changing our understanding of the disease and the way it is diagnosed. Aquaporin-4 immunoglobulin G (AQP4-IgG) is present in the majority of cases of NMOSD^1^. In 2015, the International Panel for NMO Diagnosis (IPND) decided to unify the traditional term NMO and the newer term NMOSD^2^. Furthermore, in the same year, the IPND released criteria for diagnosing NMOSD, paying regard to the clinical heterogeneity of the AQP4-IgG-negative cases and the uncertainty in diagnosis; these criteria addressed the two main categories of NMOSD: AQP4-IgG-positive and AQP4-IgG-negative, in addition to a third category where AQP4 status is unknown^2^. The core clinical characteristics of NMOSD according to the 2015 international consensus diagnostic criteria include: optic neuritis, myelitis, area postrema syndrome, narcolepsy or acute diencephalic syndrome with typical NMOSD MRI lesions, acute brainstem syndrome, and cerebral syndrome with NMOSD MRI lesions^2^.

Indeed, the immune system is at the core of the pathogenesis of NMOSD, which is essentially considered an autoimmune astrocytopathy^3^. AQP4-IgG is produced by B cells that are strongly present in the peripheral blood but rare in the CSF, which means that AQP4-IgG antibodies are mainly produced in the peripheral blood and then traverse the damaged blood-brain barrier into the brain^4^.

In pursuit of helpful diagnostic and prognostic clues and definitive pathological profiles, many studies have investigated multiple potential biomarkers of NMOSD to help establish the diagnosis and differentiate it from closely related diseases such as myelin oligodendrocyte glycoprotein antibody-associated disease (MOGAD) and multiple sclerosis (MS)^5–7^. Among these biomarkers are simple indices e.g., platelet-to-lymphocyte ratio (PLR) and cerebrospinal fluid (CSF)/serum albumin quotient (Qalb), and more specific neurological biomarkers, e.g., glial fibrillary acidic protein (GFAP) and neurofilament light (NfL)^7,8^. Simple hematological indices like platelet-to-lymphocyte ratio (PLR), neutrophil-to-lymphocyte ratio (NLR), monocyte-to-lymphocyte ratio (MLR), and systemic inflammatory ratio (SII) have been studied extensively in many autoimmune diseases^9,10^. Many studies reported associations between high NLR, PLR, and MLR and demyelinating diseases, in addition to their association with relapses^11–13^. Their implication in neuroinflammatory diseases has drawn much attention, in pursuit of any association that could be studied later for clinical relevance.

In this study, we aim to investigate whether the hematological indices, PLR and MLR, are higher in patients with NMOSD than in the healthy population, through a systematic review and meta-analysis of the available evidence.

## 2. Methods & Materials

### 2.1 Eligibility criteria

Studies should include human participants with an established diagnosis of NMOSD according to the international consensus on NMOSD 2015 criteria in addition to healthy participants. Studies should assess PLR and/or MLR in both the NMOSD group and the healthy group and provide quantitative data of MLR and/or PLR. Studies not providing quantitative data would be included in the narrative review only. Studies should be of case-control, retrospective, prospective, cross-sectional, or cohort study design.

Studies were excluded if they were in vitro studies, animal studies, conference abstracts, narrative reviews, editorials, letters without original data, or case reports. Studies were also excluded if they were not assessing MLR or PLR. Moreover, if there were duplicate reports of the same dataset, the most complete or most recent dataset was retained. If there were studies that had overlapping subjects/cohorts, all reports other than the most recent and inclusive study only were excluded.

### 2.2 Search strategy and study selection

Four databases were searched in this review: PubMed, Web of Science (WoS) Core Collection, Scopus and Cochrane CENTRAL. No language restrictions were applied. Keywords related to our study tests: “PLR”, “platelet-to-lymphocyte ratio”, “MLR” and “monocyte-to-lymphocyte ratio” and those of the emphasized condition: “NMOSD” and “Neuromyelitis Optica” were used in the search. The search started on 1 October and ended on 2 October 2026. All reports were pooled into Rayyan for screening. Search date was set from 1July 2015 to date, as accordance to IPND 2015 criteria was an inclusion criterion of our study. All duplicate studies were removed manually. Afterwards, the remaining studies were screened by 2 reviewers independently through title and abstract. When an abstract was missing, reviewers sought the abstract or full text of that report. Conflicts were resolved through discussions by the 2 reviewers.

### 2.3 Data extraction and Management

Data from each study were extracted by 2 reviewers independently. Data were extracted into preset Excel sheets. Conflicts were resolved by a third reviewer. The following data were gathered from the studies:

- Study information (e.g. Study name, design, year, country, Multi- or single center..).
- Participant characteristics (age(mean, SD), sex, number of (total participants, patients, healthy), inclusion criteria, exclusion criteria,..)
- Target condition: prevalence, severity, AQP4-IgG status, organs involved, MRI findings, active disease/remission, treatment.
- Tested Biomarkers: MLR and PLR values (mean(SD) or median(q1-q3)), Technical aspects, observer, Timing in relation to disease diagnosis and treatment if any, etc.

Studies were assessed for risk of bias using the Newcastle-Ottawa tool. Two reviewers assessed every study independently, and conflicts were resolved by a 3rd reviewer. Study authors were contacted by email if there were any gaps or missing data.

### 2.4 Data synthesis

Narrative synthesis emphasized study characteristics, disease pattern, AQP4-IgG, EDSS score and key findings. When needed, subgroups’ mean (SD) were combined into mean (SD) of the main group by repeating Cochrane’s formula. A random-effects model was selected a priori to be used for all quantitative meta-analyses due to the expected heterogeneity in clinical aspects, especially in disease activity, phenotype, and treatment status. Mean difference (MD) and 95% confidence interval (CI) were used to investigate the difference in levels of MLR and PLR between NMOSD patients and healthy subjects. This simpler method was chosen as PLR and MLR are usually calculated on the same numerical scale. For studies reporting median (1^st^ quartile-3^rd^ quartile) instead of mean (standard deviation), Wan et al. (2014)^14^ and Luo et al. (2018)^15^ methods were selected a priori to estimate the mean (SD) from the median (Q1-Q3), however, significant skewness was detected in many studies’ results. Then, we resorted to MLN method of Cai et al. (2021)^16^, yet its Box-Cox power parameter reached the upper boundary of the search interval (λ ≈ 10) in most estimates of MLR and yielded SDs smaller than the theoretical minimum (IQR/2√2). Finally, the mean and SD were estimated using the quantile estimation (QE) method of McGrath et al. (2020)^17^ through the estmeansd package of R. Between-study variance (τ²) was measured using the restricted maximum-likelihood (REML) method. Between-study heterogeneity was demonstrated by Higgins and Thomson’s I^2^ ^18^. Due to the small study number, the Hartung-Knapp^19^ adjustment was applied. Sensitivity analyses were conducted to determine the effect of sequentially removing individual studies from the analysis. Statistical analysis and graph production were done through the *meta* package on R. Subgroup meta-analyses of studies that matched patients and controls for age and sex were conducted. For (Peternell et al. 2024), Q1 and Q3 were extracted from a box plot using WebPlotDigitizer.

### 2.5 Certainty of Evidence Assessment

The certainty of evidence for the two primary outcomes, differences in monocyte-to-lymphocyte ratio (MLR) and platelet-to-lymphocyte ratio (PLR) between patients with NMOSD and healthy controls, was evaluated using the Grading of Recommendations Assessment, Development and Evaluation (GRADE) approach. Under the conventional GRADE framework, observational evidence was initially rated as low certainty. Five domains were considered for downgrading: risk of bias, inconsistency, indirectness, imprecision, and publication bias. Risk-of-bias assessments were informed by the Newcastle–Ottawa Scale; inconsistency was considered using study-specific effects, I², and prediction intervals; indirectness was assessed against the prespecified review question; and imprecision was examined using pooled 95% confidence intervals and the clinical relevance of differences. Publication bias was considered qualitatively, since fewer than ten studies were available per outcome. The resulting certainty judgments were summarized separately for MLR and PLR.

### 2.6 Protocol Registration

The protocol for this systematic review and meta-analysis was prospectively registered in the International Prospective Register of Systematic Reviews (PROSPERO; registration number: CRD420261527206).

Deviations from registered protocol: 1) Using QE to estimate mean (SD) from Median (Q1-Q3), instead of Luo et al. 2018 and Wan et al. 2014. 2) Applying the Hartung-Knapp adjustment. 3) Studies without extractable quantitative data were retained for narrative synthesis only. 4) Title change. 5) Conflicts between the findings of two reviewers were resolved by a third reviewer rather than by discussion by the two authors.

## 3. Results

Initial search gathered 146 total reports. PLR and MLR search results were inspected separately. All of these reports (titles and abstracts) were pooled into Rayyan. 70 duplicates were manually removed. Only 76 reports remained for title and abstract screening. After screening, only 11 reports were sought for full text retrieval and review. (Baradaran et al. 2025)^20^ had an English abstract and a Persian-only full text. We included the study and translated the full text with Google Translate and multiple AI tools; we could not find a Persian-speaking collaborator, so we extracted basic data from the study and did not perform a risk of bias assessment for it. It was not included in the meta-analysis. The search and screening results flow is depicted in a PRISMA flow chart (Figure 1). MLR was addressed in 7 out of 11 included studies, while PLR was investigated in all of them.

**Figure 1:**
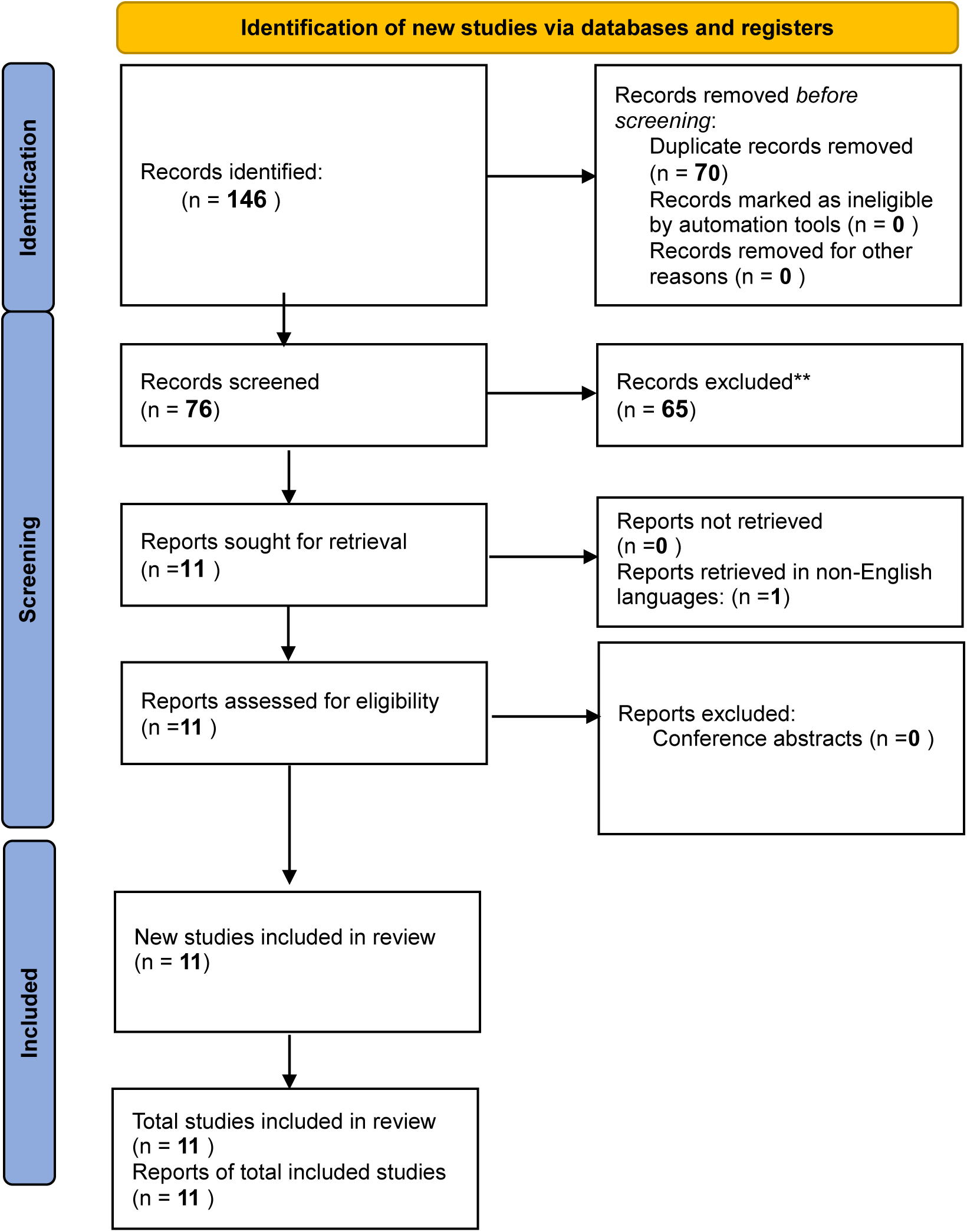
PRISMA flow diagram of search results and included studies.

All studies were single-center, except for one (Peternell et al. 2024). Eight studies were undertaken in China, and one study was conducted in each of the following countries: Austria, Japan, and Iran. All included studies enrolled patients with NMOSD according to the international consensus criteria of 2015. Five studies included both AQP4-IgG seropositive and AQP4-IgG seronegative NMOSD patients; the other 5 studies included seropositive patients only, and one study (Baradaran et al. 2025) did not mention any data on this point. Only (Peternell et al. 2024) included pediatric patients exclusively; however, more than one study stated an inclusion criterion of 14 years of age or older. All studies included patients in the acute phase, yet they enrolled patients at different time points from the start of symptoms, except for (Zhan 2026 et al.), which included 42% of patients in remission, and (Baradaran etl al. 2025), which did not emphasize disease phase. Seven studies matched controls for age and sex, while the other four did not.

Only (Wang et al. 2025) further classified NMOSD patients into disease-modifying treatment-receiving NMOSD patients (DMT-NMOSD) and (non-DMT-NMOSD). PLR in both groups, as well as in acute NMOSD, was found to be higher than in controls^21^. Interestingly, (Peternell et al. 2024) recorded PLR of patients at the acute phase, during acute treatment and at remission; the PLR value measured during the acute phase was used in the meta-analysis^22^. (Zhan et al 2026) included two distinct groups of patients: acute phase and remission groups^23^; both were combined into one before meta-analysis. It found that PLR is higher during the acute phase than during remission, in contrast to (Peternell et al. 2024) that found a higher PLR in remission than in the acute phase, which is different from MS and MOGAD, in both of which PLR was higher during the acute phase than during remission^22,23^. (Baradaran et al. 2025) found a higher PLR in NMOSD patients than in healthy controls^20^. Study characteristics are displayed in (Table 1).

**Table 1:** study characteristics. *Study design was depicted according to how studies identified themselves. Some studies reported unclear or inaccurate study design.

| **Study** | Country | Study design* | Single or Multi-center | No. of Patients (controls) | Matching for age and sex | Age(Mean, SD) of Patients | Sex | Test(s) of interest | Acute Phase/Remission (at blood sampling) | AQP4-IgG status of patients |
|---|---|---|---|---|---|---|---|---|---|---|
| Akaishi 2023<sup>13</sup> | Japan | Observational (Design not specified) | Single | 13 (65) | Yes | 47.73±14.95 | M=2 (15.4%), F=11 (84.6%) | PLR & MLR | All Patients sampled within 3 months of the last attack, before treatment | 13/13 positive (100%) |
| Fang 2023<sup>12</sup> | China | Observational (Design not specified) | Single | 54 (54) | Yes | 46.57±15.62 | M=10 (18.5%), F=44 (81.5%) | PLR & MLR | Acute phase (all patients, at admission) | 43/54 positive (79.6%) |
| Yan 2023<sup>7</sup> | China | Retrospective review of medical records | Single | 72 (72) | No | 46.74±16.23 | M=7 (9.7%), F=65 (90.3%) | PLR & MLR | Acute (disease duration <30 days) | 51/72 positive (70.8%) |
| Li 2024<sup>28</sup> | China | Retrospective study (with temporal validation cohort) | Single | 249 (249) [validation cohort: 38(85)] | Yes | 36.76±12.87 | M=69 (27.7%), F=180 (72.3%) | PLR & MLR | Acute (within 2 weeks of most recent recurrence) | 175/249 positive (70.3%) |
| Peternell 2024<sup>22</sup> | Austria | Prospective study (BIOMARKER cohort) | Multi-center | 11(39) | No | (Pediatric)<br>13 ± 3.45 (during first acute attack) | F = 10 (91%)<br>M= 1 (9%) | PLR | All patients evaluated in Acute, treatment, remission | 11/11 positive (100 %) |
| Wang 2024<sup>24</sup> | China | Observational (Design not specified) | Single | 60 (60) | Yes | 26.41±12.15 | M=30 (50%), F=30 (50%) | PLR & MLR | Acute (admitted within 2 weeks of symptom onset) | 60/60 positive (100%) |
| Wang 2025<sup>21</sup> | China | Observational (Design not specified) | Single | 76 (37) | Yes | 43.56 ± 17.19 | M=9 (11.8%)<br>F=67 (88.2%) | PLR | Acute phase, some patients were followed up | 20/20 positive (100%) |
| Sun 2025<sup>29</sup> | China | Observational (Design not specified) | Single | 79(100) | No | 48.19 ± 16.59 | M: 18 (22.8%)<br>F: 61 (77.2%) | PLR | within 1 week of new clinical symptoms or imaging findings | 59/79 positive (74.7%) |
| Zheng 2025<sup>25</sup> | China | Retrospective observational cohort study | Single | 41 (41) | No | 40.88±15.33 | M=16 (39.0%), F=25 (61.0%; paper states 50.98%) | PLR & MLR | Acute (disease duration <30 days) | 15/41 Positive (36.6%) |
| Zhan 2026<sup>23</sup> | China | Prospective cohort study | Single | 103 (25) | Yes | Acute: 44.45±12.67<br>Remission: 50.93±13.65 | M=9 (8.7%), F=94 (91.3%) | PLR & MLR | Acute phase n=60; Remission n=43 | 103/103 positive (100%) |
| Baradaran 2025<sup>20</sup>(in Persian) | Iran | Cross-sectional | Single | 75(75) | Yes | 36.11(11.47) | F=55(73.3%)<br>M=20(26.7%) | PLR | Patients with exacerbation or steroid use within 30 days excluded | Not reported |

\*

Out of 7 studies providing quantitative data of MLR in NMOSD patients and controls, (Fang et al. 2023)^12^ was disqualified from the meta-analysis due to potential overlap with the newer study (Wang et al. 2024)^24^, which included a slightly larger number of NMOSD patients. The same (Fang et al. 2023)^12^ was disqualified from the meta-analysis of PLR due to the same reason, in addition to (Wang et al. 2025)^21^, since it did not provide quantitative data, and (Baradaran et al. 2025), as it did provide median and IQR without 1^st^ and 3^rd^ quartiles. The screening concluded with the inclusion of only 8 studies out of 11 for PLR meta-analysis.

For MLR, the total number of NMOSD patients included in studies eligible for meta-analysis was 538 patients in addition to 512 healthy controls. For PLR, studies that qualified for meta-analysis included 628 patients and 651 healthy controls.

For MLR, random-effects meta-analysis yielded 0.125 mean difference (MD) with a 95% confidence interval (CI) of [-0.114; 0.363]. The prediction interval was [-0.3 72; 0.621]. Heterogeneity was high among studies investigating MLR, (I² = 83.8%, 95% CI: 66.2%–92.2%), and τ² was 0.0315 (Figure 2). Leave-one-out sensitivity analysis revealed no substantial change in MD or heterogeneity, except when (Zhen g et al. 2025) was omitted. Leaving Zheng et al. (2025) out resulted in a significant change in MD [95% CI] from 0.125 [-0.114; 0.363] to 0.06 [0.02 – 0.10], and I^2^ went down to 69.2%. (Figure 4)

**Figure 2:**
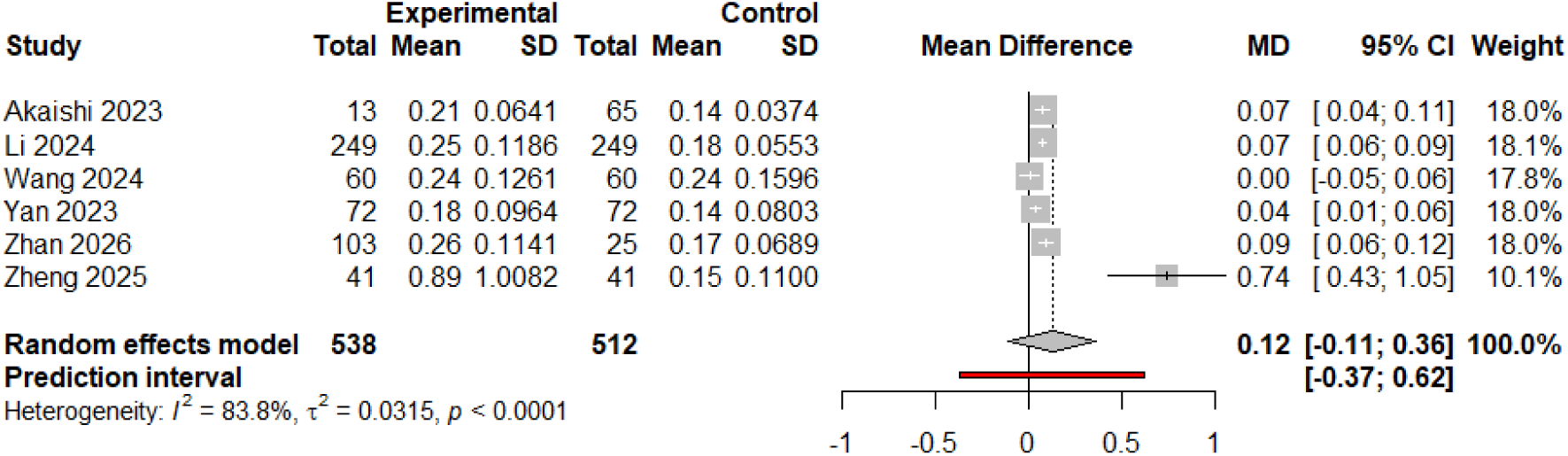
MLR main analysis results.

Subgroup meta-analysis of studies that carried out matching for age and sex (n=4) demonstrated a pooled MD of 0.066 with a 95% CI of [0.012; 0.12]. The prediction interval was [-0.026; 0.158]. Regarding heterogeneity, I^2^ was 61.9% [0.0%; 87.2 %]; while τ² equaled 0.0006. (Figure 3)

**Figure 3:**
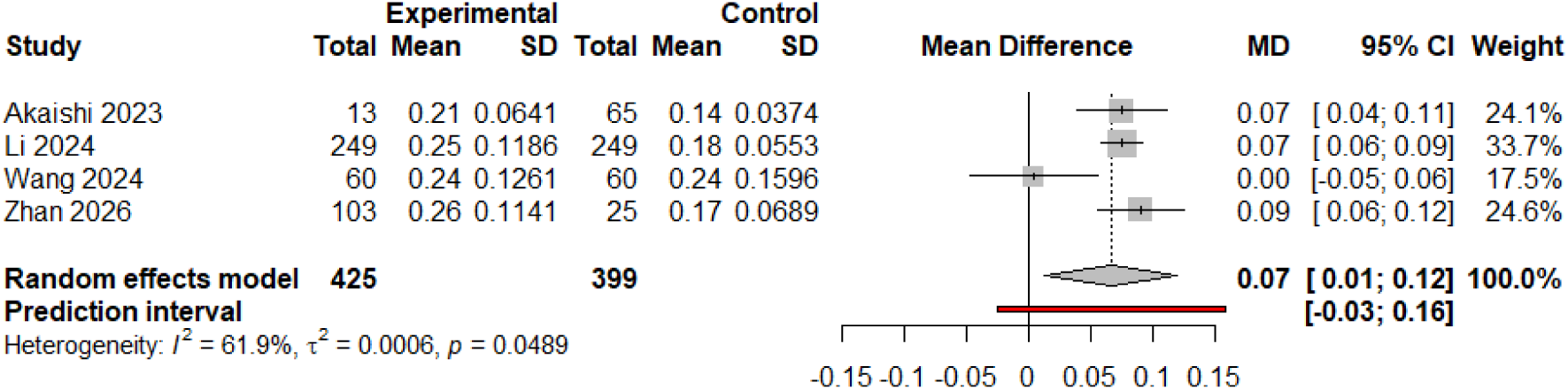
Results of MLR Subgroup analysis of age- and sex-matched studies.

**Figure 4:**
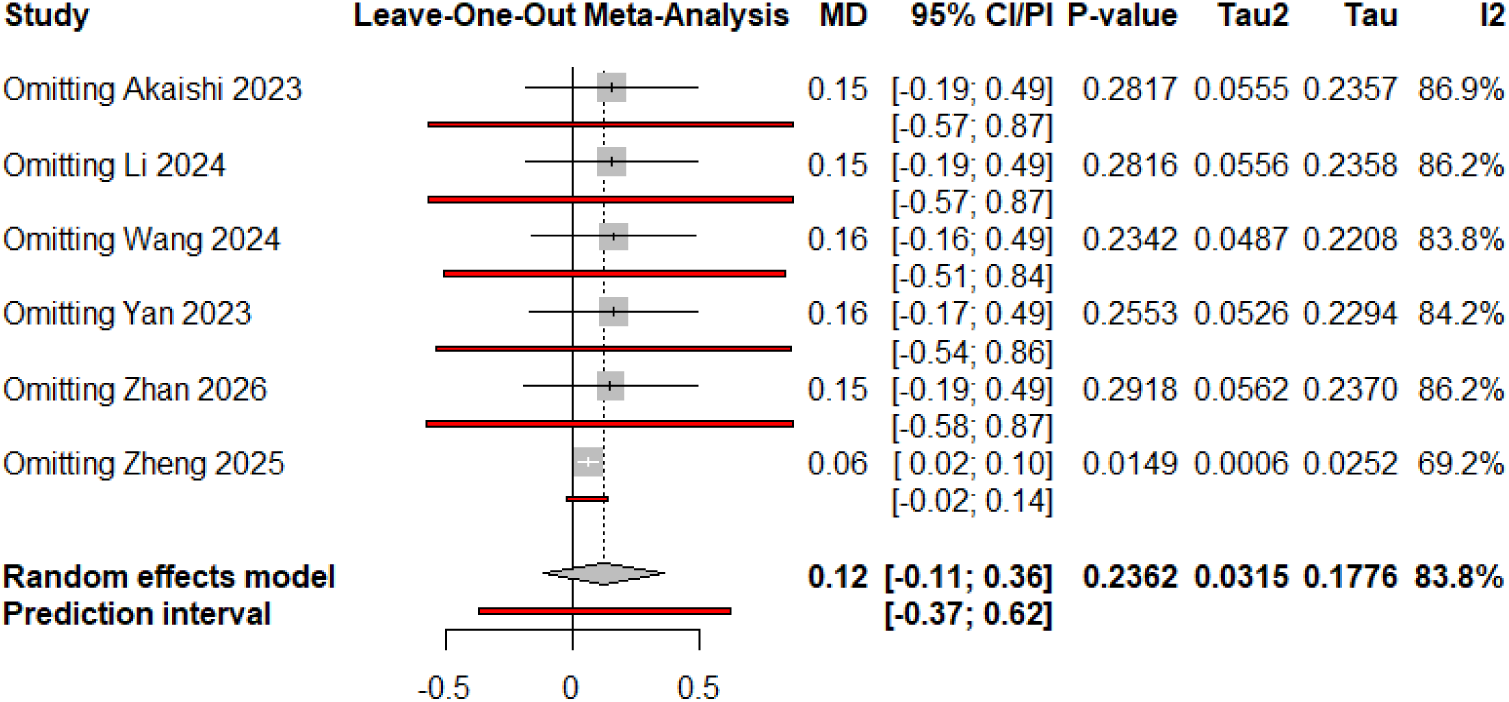
MLR leave-one-out sensitivity analysis results.

For PLR, random-effects meta-analysis revealed a pooled mean difference (MD) of 23.99 with a 95% confidence interval of [6.03; 41.96]. Heterogeneity between studies was significantly high; I^2^ was 82.3% [66.3%; 90.7%]; and the figure for τ² was 339.61. The 95% prediction interval was [-23.3; 71.29]. Leave-one-out sensitivity analysis revealed that the effect size and heterogeneity were generally stable, except for (Zheng et al 2025), which when excluded, the estimated pooled MD dropped to 17.85 [1.2 – 34.49] and I^2^ fell to 73.5%. In contrast, omitting (Li et al. 2024) lead to an increase in the pooled MD estimate to 28.97 [11.77; 46.18] while I^2^ fell to 63.6%. (Figure 5, 7)

**Figure 5:**
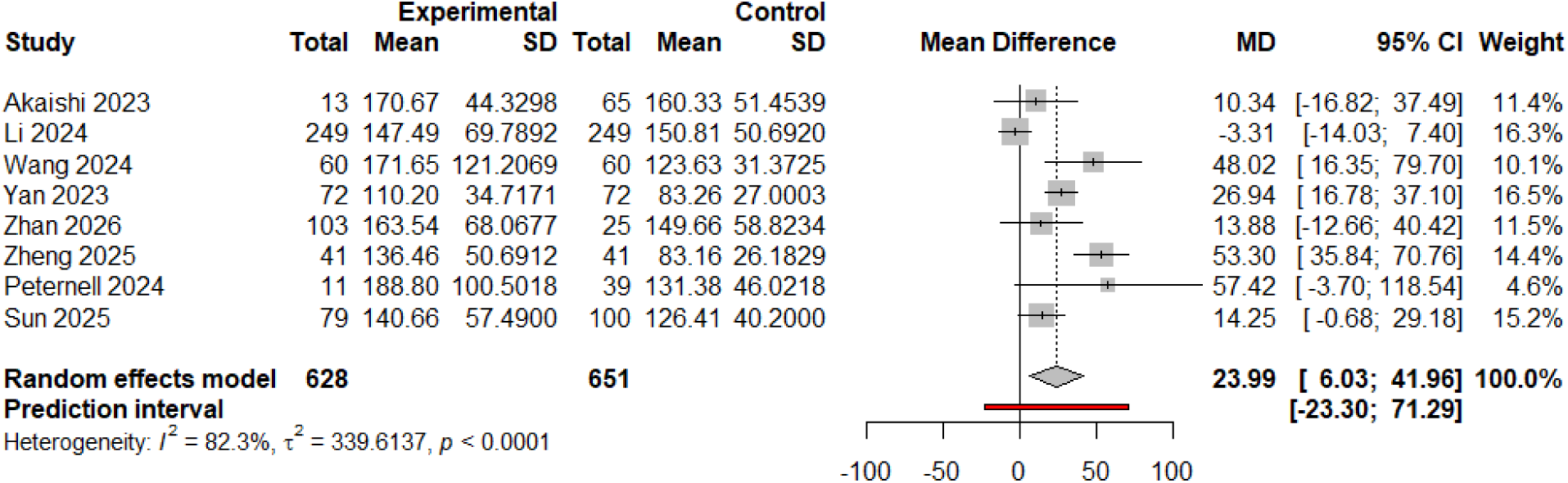
PLR main analysis results.

Subgroup analysis for studies that complied with matching patients and controls for age and sex for PLR (n=4), showed a pooled MD [95% CI] of 14 [-19.68; 47.68]. The prediction interval was [-49.98; 77.98]. I^2^ was 69.8%, and τ² was 293.69. (Figure 6)

**Figure 6:**
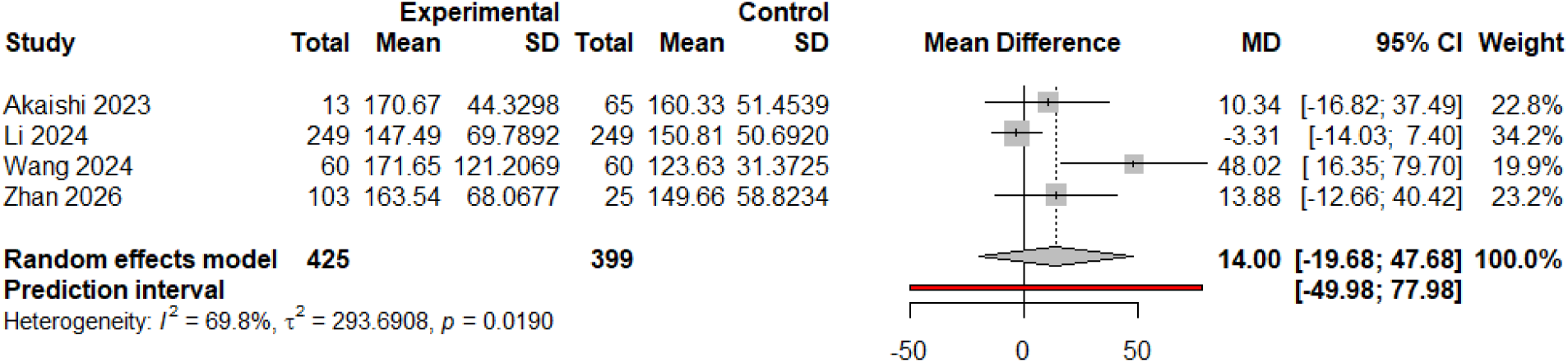
Results of PLR Subgroup analysis of age- and sex-matched studies.

**Figure 7:**
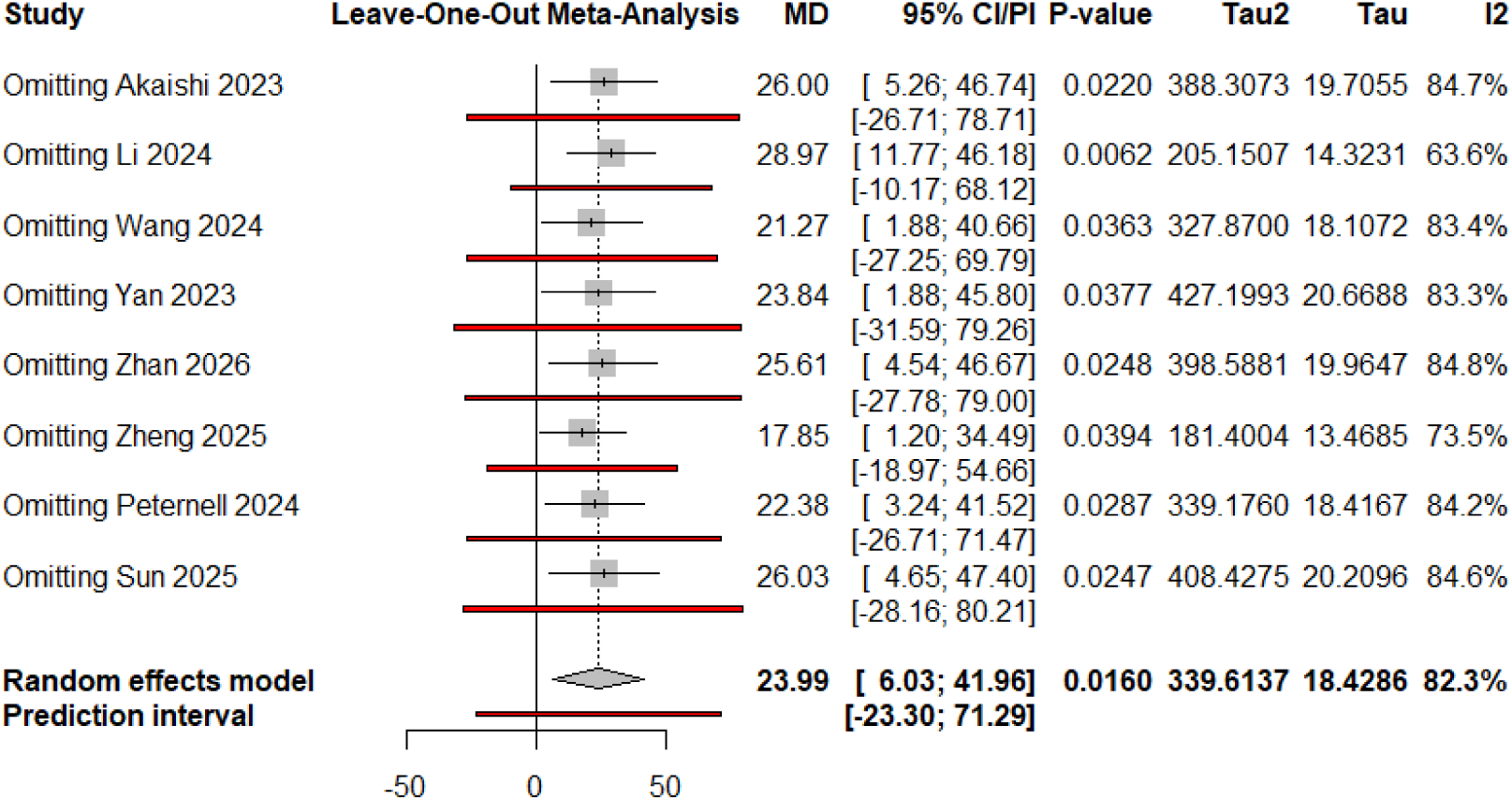
PLR leave-one-out sensitivity analysis results.

Risk of bias assessment of studies yielded a fair rating of 4-6 out of 9 for six studies, a poor rating of 0-3 out of 9 for two studies, and a good rating of 7 out of 9 for two studies. (Table 2)

**Table 2:** NOS quality assessment results.

| **Study** | **Study Direction** | **Selection (max 4)** |  |  |  | **Comparability (max 2)** |  | **Exposure (max 3)** |  |  | **Total stars (max 9)** | **Quality** |
|---|---|---|---|---|---|---|---|---|---|---|---|---|
|  |  | **Adequate case definition** | **Representativeness of the Cases** | **Selection of controls** | **Definition of controls** | **Controls for age & sex** | **Controls for additional factor ( inflammatory conditions)** | **Exposure ascertainment** | **Same method for cases & controls** | **Non-response rate** |  |  |
| **Akaishi 2023** | Unclear | ★ |  | ★ |  | Yes ★ | No | ★ | ★ | NA | 5 | **Fair** |
| **Fang 2023\*** | Retrospective + prospective follow-up of some participants | ★ |  |  |  | Yes★ | No | ★ | ★ |  | 4 | **Fair** |
| **Yan 2023** | Retrospective | ★ |  |  |  | No | No | ★ | ★ | NA | 3 | **Poor** |
| **Li 2024** | Retrospective + temporal validation | ★ |  |  | ★ | Yes ★ | Yes ★ | ★ | ★ | ★ | 7 | **Good** |
| **Peternell 2024\*\*** | Prospective | ★ |  | ★ |  | NO | Yes ★ | ★ | ★ |  | 5 | **Fair** |
| **Wang 2024** | Unclear | ★ |  | ★ |  | Yes ★ | No | ★ | ★ | NA | 5 | **Fair** |
| **Wang 2025\*** | Prospective | ★ |  |  | ★ | Yes ★ | Yes ★ | ★ | ★ | ★ | 7 | **Good** |
| **Sun 2025** | Retrospective | ★ |  |  | ★ | No | Yes ★ | ★ | ★ | NA | 5 | **Fair** |
| **Zheng 2025** | Retrospective | ★ |  |  |  | No | No | ★ |  | NA | 2 | **Poor** |
| **Zhan 2026** | Prospective | ★ | ★ |  |  | Yes ★ | No | ★ | ★ | ★ | 6 | **Fair** |
| **Baradaran 2025\*** |  |  |  |  |  |  |  |  |  |  |  |  |

★: star awarded. Total out of 9 stars. Quality: Good = 7–9, Fair = 4–6, Poor = 0–3.
\* Study NOT enrolled in meta-analysis.
\*\* Peternell 2024 was assessed with the NOS cohort form; its items respectively correspond to the columns as follows: representativeness of the exposed cohort; selection of the non-exposed cohort; ascertainment of exposure; outcome not present at start of study; comparability for age & sex; comparability for additional factors; assessment of outcome; follow-up long enough; adequacy of follow-up.
Study direction: classification of each study as prospective or retrospective as reported/judged by the reviewers; "Unclear" = not clearly stated in the paper.
The non-response rate is non-applicable for studies of retrospective or unclear direction and hence were given NA in this field; this should be taken into account when considering risk of bias of these studies.

The certainty of evidence was rated very low for both outcomes (Table 3). For MLR, the pooled mean difference was 0.125 (95% CI −0.114 to 0.363; six studies; 1,050 participants), with substantial between-study heterogeneity (I² = 83.8%). For PLR, the pooled mean difference was 23.99 (95% CI 6.03 to 41.96; eight studies; 1,279 participants), also with substantial heterogeneity (I² = 82.3%). Both bodies of evidence were downgraded for methodological limitations and unexplained inconsistency. MLR was additionally affected by imprecision because its confidence interval spanned both lower and higher values in NMOSD. No downgrading for indirectness was considered necessary. Publication bias remained undetermined. Accordingly, confidence in the magnitude and reproducibility of the observed differences is limited.

**Table 3.** GRADE Summary of Findings.

| **GRADE assessment** | **MLR** | **PLR** |
|---|---|---|
| Number of studies | 6 | 8 |
| Participants (NMOSD / healthy controls) | 538 / 512 | 628 / 651 |
| Pooled mean difference (95% CI) | 0.125 (−0.114 to 0.363) | 23.99 (6.03 to 41.96) |
| Risk of bias | Serious<sup>a</sup> | Serious<sup>a</sup> |
| Inconsistency | Serious<sup>b</sup> | Serious<sup>b</sup> |
| Indirectness | Not serious | Not serious |
| Imprecision | Serious<sup>c</sup> | Serious<sup>d</sup> |
| Publication bias | Undetermined<sup>e</sup> | Undetermined<sup>e</sup> |
| **Overall certainty of evidence** | **Very low** | **Very low** |

**a. Risk of bias: Downgraded because of methodological limitations identified through the Newcastle–Ottawa Scale, including concerns regarding participant selection, comparability between groups, and residual confounding.**
**b. Inconsistency: Downgraded because of substantial unexplained between-study heterogeneity (MLR: I² = 83.8%; PLR: I² = 82.3%), with prediction intervals encompassing the null value.**
**c. MLR imprecision: Downgraded because the pooled 95% confidence interval crossed the null value and included differences in both directions, limiting confidence in the direction and magnitude of the association.**
**d. PLR imprecision: Downgraded because, although the pooled 95% confidence interval excluded the null value, it encompassed a broad range of effect magnitudes (MD 6.03–41.96). In the absence of an established clinically meaningful difference for PLR, uncertainty remains regarding the clinical importance of the observed association.**
**e. Publication bias: Could not be reliably evaluated because fewer than ten studies were included in each meta-analysis. No additional downgrading was applied.**
**Abbreviations: CI, confidence interval; GRADE, Grading of Recommendations Assessment, Development and Evaluation; MD, mean difference; MLR, monocyte-to-lymphocyte ratio; NMOSD, neuromyelitis optica spectrum disorder; PLR, platelet-to-lymphocyte ratio.**
**Table note: Certainty was evaluated using the conventional GRADE framework, with observational evidence initially rated as low. Final certainty was very low for both outcomes because of risk-of-bias concerns and substantial unexplained heterogeneity. Publication bias was not formally assessed.**

## 4. Discussion

Our analysis of studies investigating MLR produced mixed results. Although the main analysis resulted in a mean difference of 0.125, the 95% confidence interval [-0.114; 0.363] crossed zero, meaning the pooled effect estimate is statistically non-significant. In addition, the I^2^ is significantly high. Furthermore, (Zheng et al. 2025) reported a substantially larger effect (MD = 0.74)^25^. Leave-one-out sensitivity analysis omitting (Zheng et al. 2025) lead to a decrease in the pooled MD [95% CI] to 0.06 [0.02 – 0.10]; in addition, I^2^ decreased from 83.8% to 69.2%.

This suggests that the results of the main analysis should be interpreted cautiously, as results are highly sensitive to one study (Zheng et al. 2025), which was not among studies that matched controls for age and sex. Furthermore, the subgroup analysis of studies that matched NMOSD patients and controls for age and sex, which did not include (Zheng et al. 2025), resulted in a pooled MD [95% CI] of 0.066 [0.012; 0.12], but with a zero-crossing 95% prediction interval of [-0.026; 0.158]. This means that the true effect in a future study could be zero or even slightly in the opposite direction despite the statistically significant pooled effect estimate.

Despite evidence suggesting an association of higher MLR in NMOSD patients than in healthy subjects, the existence and magnitude of increase remain uncertain due to the limitedness and inconsistency of the available evidence. Further research would be needed to rule in or out a higher MLR association with NMOSD.

Regarding PLR, the main meta-analysis revealed a relatively modest pooled mean difference (MD) of 23.99 with a 95% confidence interval of [6.03; 41.96]. The prediction interval was [-23.3; 71.29]; meaning the true effect size in a future study could be zero or negative. Subgroup meta-analysis of studies matching patients and controls for age and sex produced a smaller pooled MD [95% CI] of 14 [-19.68; 47.68], which is statistically non-significant as the 95% CI includes zero. Omitting (Zheng et al. 2025) from the analysis yielded a smaller pooled effect size (MD) of 17.85, which was closer to the result of the analysis of age- and sex-matched studies.

(Zheng et al. 2025) displayed an outlier effect in MLR analysis and to a lesser extent, in PLR analysis. This, in addition to its high risk of bias, should be considered when interpreting the results of analyses.

In terms of clinical applicability of such results, a PLR pooled MD of 23.99 is statistically significant; however, it does not mean that it can provide any practical advantage in everyday clinical practice, because PLR has its physiological variability, and its normal range is wide and highly affected by age and sex. A 2026 study estimated the normal range of PLR to be 70-231 in females and 61-183 in males^26^. An MD increase of 23.99 might be questionable in terms of clinical value due to this wide range. It is noteworthy that the patients’ and controls’ median (Q1-Q3) figures of PLR across most included studies were within the said normal ranges. (Moosmann et al. 2021) warned that ignoring reference intervals of biomarkers when comparing cases and controls makes it difficult to interpret their clinical significance^27^.

There are several limitations to our study. First, the studies measured PLR and MLR at different time points from the first/most recent clinical attack, and some included patients in remission. This may contribute to heterogeneity. Second, most included studies were conducted in China, which may limit the generalizability of findings. Third, many of the included studies are retrospective with a significant risk of bias. Fourth, most PLR mean (SD) values were estimated from median (Q1-Q3). Fifth, there was an included study (investigating PLR) in Persian. However, a trusted human translation was not at our disposal, and since lack of accurate translation might risk the accuracy of data extraction and quality assessment, we only did basic data extraction and no quality assessment was done for the study. The study did not mention mean (SD) and stated the median (IQR) only, without specifying Q1and Q3, which meant it was not qualified for meta-analysis. Sixth, many included studies lacked clear and definitive reporting of study design; many identified as ‘retrospective’ with no well-corroborated methods section or no clearly stated design. Some studies, such as Akaishi et al. (2023), had properties of both cross-sectional and case-control studies, as they used a case-control sampling method and a snapshot-like measurement of indices. We reported the stated designs in their manuscripts. For the risk-of-bias assessment, we opted to consider those studies enrolling patients and controls for PLR or MLR assessment as case-control, as assessing control selection is crucial to our study. In addition, the ‘non-response rate’ question of NOS is not applicable to retrospective studies; hence, retrospective studies were not given a star for this point. Although it didn’t affect the overall assessment results except for one study, this should be taken into consideration. The risk-of-bias assessment revealed concerns of bias in multiple studies; Most scores (6 out of 10) ranged from 4 to 6 out of 9, meaning a moderate risk of bias, and two studies had a high risk of bias. Only two out of ten studies achieved a good score, which means a low risk of bias. Finally, the GRADE assessment indicated very low certainty of evidence for both MLR and PLR comparisons, principally because of methodological limitations and substantial unexplained between-study heterogeneity. For MLR, the pooled estimate was additionally imprecise, with a confidence interval spanning both lower and higher biomarker values in NMOSD. Although the PLR meta-analysis yielded a statistically significant pooled difference, uncertainty regarding reproducibility and clinical significance persists. The PLR pooled estimate was also deemed imprecise as it encompassed a broad range of effect magnitudes. These findings should therefore be interpreted cautiously and should not be regarded as evidence of independent diagnostic utility.

## 5. Conclusion

The main analysis suggests that MLR is not higher in NMOSD patients than in healthy controls. However, both the subgroup analysis of age- and sex-matched studies and the leave-one-out analysis excluding the outlier study showed a statistically significant MLR increase in patients. PLR was found to exhibit a modest increase in NMOSD patients compared with healthy controls. Nonetheless, the subgroup analysis of age- and sex-matched studies showed a statistically non-significant increase. The results for should be interpreted cautiously. The available evidence on both PLR and MLR is inconsistent, and further research is needed.

## Data Availability

All data produced in the present work are contained in the manuscript

## List of abbreviations

AQP4-IgG: Aquaporin-4 immunoglobulin G
CI: Confidence interval
CSF: Cerebrospinal fluid
EDSS: Expanded Disability Status Scale
GRADE: Grading of Recommendations Assessment, Development and Evaluation
IPND: International Panel for NMO Diagnosis
LETM: Longitudinally extensive transverse myelitis
MD: Mean difference
MLR: Monocyte-to-lymphocyte ratio
MOGAD: Myelin oligodendrocyte glycoprotein antibody-associated disease
MS: Multiple sclerosis
NLR: Neutrophil-to-lymphocyte ratio
NMO: Neuromyelitis optica
NMOSD: Neuromyelitis optica spectrum disorder
NOS: Newcastle–Ottawa Scale
PLR: Platelet-to-lymphocyte ratio
PRISMA: Preferred Reporting Items for Systematic Reviews and Meta-Analyses
Q1: First quartile
Q3: Third quartile
REML: Restricted maximum likelihood
SD: Standard deviation
SII: Systemic inflammatory index
WoS: Web of Science

## Declarations

- Ethics approval and consent to participate: Not applicable
- Consent for publication: Not applicable
- Availability of data and materials: All study data are available upon request.
- Competing interests: Authors declare no conflict of interest.
- Funding: This study had no external funding
- Authors’ contributions:

**Conceptualization:** Abdurrahman Elsayed Shouman

**Methodology:** Abdurrahman Elsayed Shouman

**Literature search:** Abdurrahman Elsayed Shouman

**Screening and study selection:** Abdurrahman Elsayed Shouman, Mohammad Bdour

**Data extraction:** Mahmoud Sherif, Hossam Rostom, Reem Saad, Mohammad Bdour, Asaad Ibrahim

**Risk of bias assessment:** Mahmoud Sherif, Hossam Rostom, Reem Saad, Mohammad Bdour, Asaad Ibrahim, Abdurrahman Elsayed Shouman.

**Formal analysis / Meta-analysis:** Abdurrahman Elsayed Shouman

**Software:** Abdurrahman Elsayed Shouman

**Certainty of Evidence assessment (Methodology & Writing)**: Reem Saad

**Writing – original draft:** Abdurrahman Elsayed Shouman

